# Early intervention with a glycerol throat spray containing cold-adapted cod trypsin after self-diagnosis of the common cold: a randomised prospective, parallel group and single-blind methods trial (Glycerol-cod trypsin spray in common cold)

**DOI:** 10.1101/2021.10.11.21264744

**Authors:** B. Fredrik Lindberg, Ida Nelson, Jonas Ranstam, Donald K. Riker

## Abstract

**Background:** A glycerol throat spray containing cold-adapted cod trypsin (GCTS) deactivates common cold virus *in vitro* and decreases pharyngeal rhinovirus load after inoculation in humans. We relied on early self-diagnosis and evaluated two different scales to detect a treatment effect in naturally occurring common colds.

**Methods:** Adults were enrolled in this randomised, prospective, parallel group, single-blind study to begin treatment six times daily at first sign of a common cold or were assigned to a non-treated group. Jackson’s symptom scale and the 9-item Wisconsin Upper Respiratory Symptom Survey (WURSS)-21 quality of life (QoL) domain were recorded daily by subjects and area under the curve over 12 days (AUC_1-12_) calculated.

**Results:** Treatment resulted in reduced symptoms with an AUC_1-12_ of 45.1 ± 32.5 for Jackson scores compared to 53.8 ± 35.7 in the controls (p=0.023). AUC_1-12_ for the 9-item WURSS-21 QoL domain was likewise improved, 113.6 ± 107.7 and 152.7 ± 126.3 (p=0.006), respectively. During the first four days fewer of the treated subjects (35.3%) used rescue medication than did the control group (50.4%, p=0.014).

**Conclusions:** Reduction in common cold symptoms was seen with treatment with a glycerol throat spray containing cold-adapted cod trypsin. This effect was best detected with the 9-item WURSS-21 QoL domain.

## BACKGROUND

The common cold is a self-limiting respiratory viral infection not only affecting the individual, but also the society in its high costs and lost productivity [1, 2]. The cause of the common cold is infection by one of over 200 known respiratory viruses, typically rhinoviruses, or coronaviruses, influenza viruses, adenoviruses, parainfluenza viruses, respiratory syncytial viruses and enteroviruses [3]. Rhinovirus is well adapted to its host initially overcoming innate epithelial barriers, interferon release and adaptive immune responses. The diversity of respiratory viral pathogens has so far foiled any attempt to find a universal cure [4–6]. After inoculation virus is transported from the nose back to the nasopharynx where infection of the mucosa is first established [7]. Local symptom development starts with sore scratchy throat and malaise quickly followed by nasal congestion, rhinorrhoea, sneezing and finally cough [8].

In an attempt to halt local virus propagation during the incubation period and thereby alleviating and shortening the common cold, a technology has been developed consisting of a hyperosmotic glycerol solution containing cold-adapted trypsin from the Atlantic cod (*Gadus morhua*) (GCTS) [9]. When sprayed onto the throat the solution forms a transient barrier across the oro-pharynx that acts osmotically while at the same time deactivating proteins viruses use to bind and enter the host cell. The spray solution has demonstrated broad antiviral activity *in vitro* deactivating 64-100% of virus activity [10, 11]. In a double-blind pilot study of healthy volunteers inoculated intranasally with rhinovirus-16 the barrier solution reduced pharyngeal virus load significantly compared to placebo and reduced the disease progression assessed by the Jackson scale [12].

In order to intervene at the very first sign of a common cold we conducted a prospective, single-blind, randomised and comparative multicentre methodology study to investigate a clinical design that relies on self-diagnosis to start treatment at the first perceived signals of common cold presence. We believed that early and sustained intervention during the prodromal phase would reduce viral infectivity, replication and local spread resulting in a reduced symptom burden, improved quality of life and possibly fewer days of colds illness. The primary objective of this study was to investigate the ability of two different rating scales to detect these meaningful benefits of early intervention in the common cold with GCTS.

## METHODS

### Study design and population

This was a prospective, multicentre, randomised, single-blind (investigator) study using a parallel-group design that compares early intervention with the investigational device (GCTS) to no treatment in patients with naturally occurring common colds. After local Ethics Committee approval at Charité Hospital (Berlin, Germany), and written informed consent, subjects were recruited to six centres located in the Berlin area through advertising. Women of child-bearing potential with a negative pregnancy test and agreeing to use appropriate contraception methods were eligible for inclusion. Men and women (18-70 years) with at least three occurrences of common cold within the last 12□months, but generally in good health and ready to comply with trial procedures were eligible and screened for entry against inclusion and exclusion criteria. Exclusion criteria consisted of known allergy or hypersensitivity to the components of the investigational product, history and/or presence of a clinically significant condition/disorder which per investigator’s judgement could interfere with the results of the study, or the safety of the subject, influenza vaccination within the last 3 months prior to V1 and during the study, regular use of products that could influence the study outcome, pregnant or nursing women, history of or current abuse of drugs, alcohol or medication, participation in the present study of a person living in the same household as the subject, inability to comply with study requirements according to investigator’s judgement and participation in another clinical study within 30 days of V1 or during the study.

There were no changes to the study design during the conduct of the study. In addition to the analyses defined in the statistical analysis plan (SAP) post hoc analyses were performed as defined in the addendum to the SAP.

The trial was registered at ClincialTrials.gov, number NCT03831763, and was performed in compliance with the principles of the World Medical Association (Declaration of Helsinki), ICH GCP E6, German Act on Medical Devices (MPG) §23b and ISO 14155.

### Randomisation and masking

The subjects, the study site staff handling the investigational product, and the study monitors were aware of the allocated arms. All others involved in the study, including investigators performing the clinical assessments of common cold, statisticians and the sponsor were blinded to the allocation. The subjects were randomised (1:1) at the screening visit to the treatment or control group. Randomisation numbers were assigned to the subjects in a sequential order upon enrolment by the investigator, based on time of randomisation at each investigational site. Several whole blocks of four were allocated to each site. The randomisation list was concealed and stored under lock and key by the sponsor until after database closure and sign-off of the statistical analysis plan at which time it was provided to the external statistician responsible for the statistical analysis.

### Procedures

At the screening and enrolment visit eligible subjects once randomised (1:1) to one of two groups received the subject diary to be used throughout the study. Those randomised to the treatment group received the investigational product, a throat spray consisting of glycerol, water, trypsin secreted from the pyloric caeca of the Atlantic cod (*Gadus morhua*), ethanol (<1 %), calcium chloride, trometamol (Tris buffer) and menthol (ColdZyme^®^ Mouth Spray, Enzymatica, Sweden) to be sprayed twice (1 dose = 340 µl) every second hour up to 6 times daily. The subjects were blinded to the commercial name as the investigational products were relabelled for investigational use only. All participants were instructed to start the diary ratings at the start of a cold when answering “Yes” to the binary question “Do you think you might be having the first signs of a common cold?” and simultaneously experiencing a total Jackson score of at least 1 (mild = present, but not disturbing or irritating) for any symptom except headache. Subjects in the treatment arm of the study were instructed to start treatment at that time. It is unlikely that the allocation process at the initial enrolment visit during which subjects were entered into the colds surveillance period would introduce investigator bias affecting randomly occurring colds at a later time in either group.

From the first day of entering the treatment phase the subjects started to record their symptoms twice daily once in the morning and once in the evening using the Jackson scale [13] and once daily in the evening using the 9-item Wisconsin Upper Respiratory Symptom Survey (WURSS-21) Quality of Life (QoL) domain [14]. All subjects answered a daily binary question in the evening “Do you think that you are still having a common cold?”. They also recorded the use of the investigational product (treatment group only) and any use of rescue medication, including paracetamol (acetaminophen), maximum 2□g/day, ibuprofen, maximum 400 mg/day or saline nose drops or spray, if needed for symptom relief.

The Jackson score was calculated by summing the scores for the following 8□symptoms: sore throat, blocked nose, runny nose, cough and sneezing (local symptoms) as well as headache, malaise, and chilliness (systemic symptoms). Symptoms were assessed on a 4-point scale: 0□= □none, 1□= □mild, 2□=□moderate, 3□= □severe. The 9 item (items 12 to 20) quality of life (QoL) domain of the WURSS-21 was calculated by summing the individual scores recorded for the question “How much has your cold interfered with your ability to…: Think clearly, sleep well, breathe easily, walk-climb stairs-exercise, accomplish daily activities, work outside the home, work inside the home, interact with others, and live your personal life”. Response options ranged from 0 = “Not at all” to 7 = “Severely”. A lower value corresponds to an improved quality of life.

Within 3 days of experiencing the first perceptions of a common cold and having had recorded subjective symptoms the subjects presented to the investigator for a physical examination and confirmation of common cold signs. The investigator on site was blinded as to a subject’s allocation. The third and last visit to the study site had to take place by day 16 (+/−4 days) after the start of a cold. Subjects with no symptoms during the study period attended a termination visit 16 weeks (+/−7 days) after enrolment. These terminal visits would not influence subjective data already collected.

### Outcomes

Primary outcomes were individual daily ratings using the Jackson scale and the 9-item QoL domain subscore taken from the Wisconsin Upper Respiratory Symptom Survey WURSS-21. Using these data area under the curve was calculated over the 12 days from the start of a cold (AUC_1-12_). AUC is a measure of the total symptom burden suffered during a common cold.

Other outcomes included: investigator confirmation of participants’ self-diagnosis through objective signs within 3 days after symptom start, use of concomitant symptom relieving rescue medication, adverse events, device deficiencies, duration of common cold, and global evaluation of efficacy. Measures of duration of a cold do not reflect the severity, symptom burden or bothersomeness of the illness, only a return to absolute wellness. Primary data in case record forms and subject diaries were used to assess endpoints.

### Statistical Analyses

The number of subjects enrolled was determined on the basis of a previous pilot study [12] and the inter- and intra-individual variability collected from other published studies [15, 16]. The enrolment was extended from the planned 300 subjects to include a total of 400 subjects during the course of the study, to ensure entry of a minimum of 200 subjects experiencing common cold symptoms.

The full analysis set (FAS) consists of all randomised subjects meeting the inclusion/ exclusion criteria (n=400) and for whom there is at least one observation after the onset of common cold symptoms. Subjects in the treatment group must have received at least one dose of the investigational product (n=273) excluding those who at a later stage were detected to have violated the exclusion/inclusion criteria (n=267, treated=136 and control group=131).

Safety analyses included all randomly assigned participants. The definition of the FAS population took place prior to unblinding and statistical analysis.

All group comparisons are exploratory. Unless otherwise stated the evaluation of endpoints is based on the FAS. Individual areas under the curve (AUC) for Jackson scores and the 9-item WURSS-21 QoL domain were assessed by applying the trapezoidal approximation over 12 days and presented as means and SD. Hypothesis-generating statistical tests comparing the two groups were performed using the Mann-Whitney-U test. The Wilcoxon’s signed ranks test was used for pre/post comparisons within the treatment groups, and the χ2-squared test is used for comparing frequencies between the two treatment groups. Individual items of the WURSS-21 and Jackson scores were compared between the treatment groups using normal deviate tests (z-tests) of the mean AUCs. The results are presented in a forest plot produced by the package forest plot with R version 3.5.2. The statistical calculations were performed using SPSS Statistics version 22.0.

## RESULTS

Between Jan 25, 2018, and April 20, 2018, 400 eligible subjects were recruited and randomly assigned to treatment group (n=200) or control group (n=200). The last patient was completed on June 5, 2018. After perceiving the start of a common cold, subjects entered the treatment phase (n=273, Figure 1). Prior to unblinding protocol deviations were identified and classified resulting in six exclusions from the FAS (three from each group). Baseline characteristics are described in Table 1.

**Table 1:** Baseline characteristics of the full analysis set (FAS) population.

|  | Treatment group<br>(n=136) | Control group<br>(n=131) | *Table 1: Baseline*<br>*characteristics of the full*<br>*analysis set (FAS)*<br>*population.* |
| --- | --- | --- | --- |
| Age (years) | 42.7 (13.8) | 38.3 (13.2) |  |
| Sex (female) | 92 (67.6 %) | 98 (74.8 %) |  |
| Caucasian | 132 (97.1 %) | 127 (96.9 %) |  |
| Systolic blood pressure (mm Hg) | 126.6 (13.8) | 124.8 (12.1) |  |
| Diastolic blood pressure (mm Hg) | 78.2 (7.9) | 77.0 (7.6) |  |
| Pulse rate (bpm) | 73.2 (7.9) | 72.0 (8.6) |  |

Data are mean (SD); n (%).

**Figure 1:**
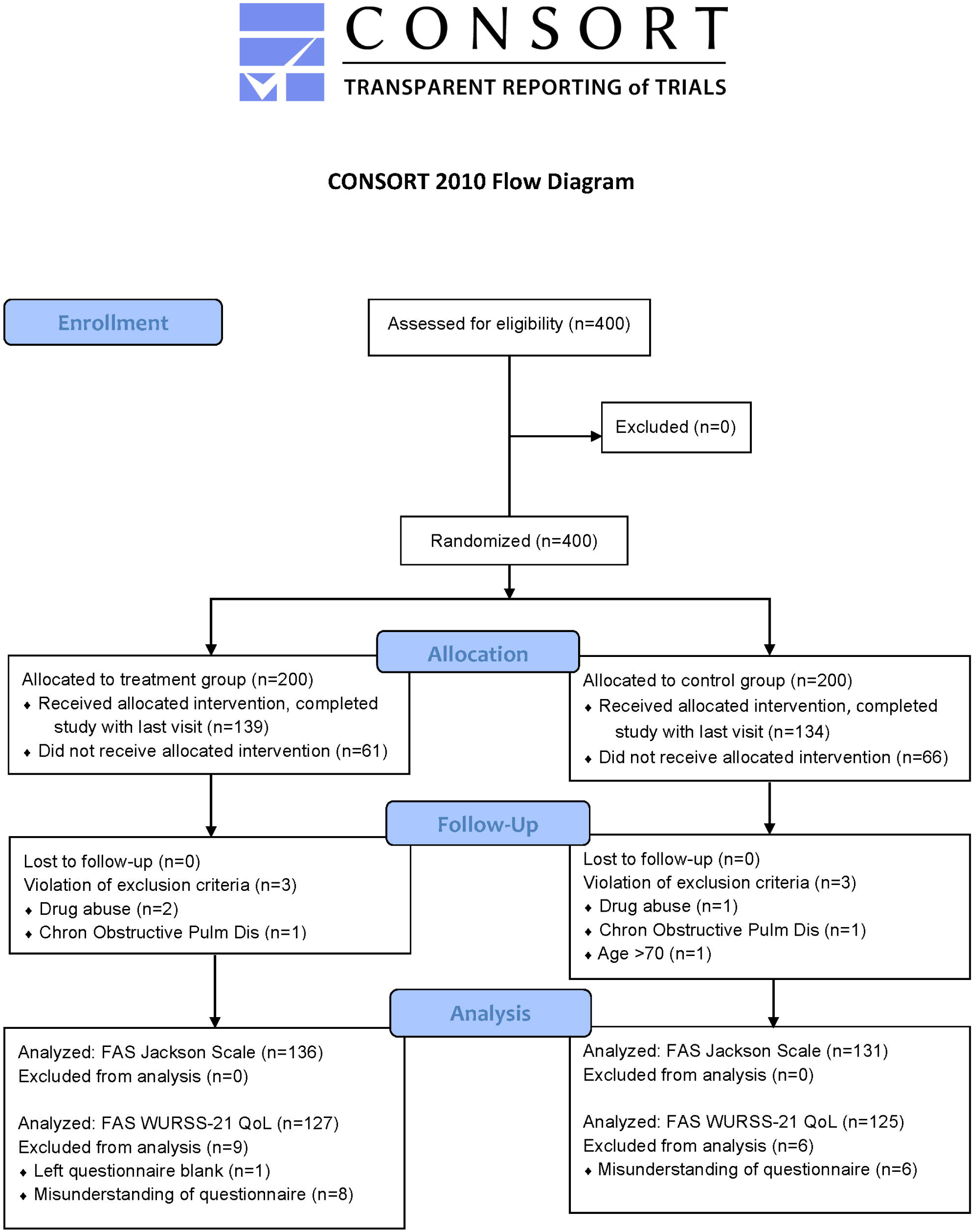
Trial profile.

The common cold started with an almost identical burden of symptoms in the two groups with a Jackson score of 7.59 ± 4.68 in the treatment group and 7.46 ± 5.34 in the control group (p=0.654) on the first morning. The signs of a common cold was confirmed clinically by the blinded investigator within three days from the start of a perceived common cold for all subjects in the control group (131 of 131), and for 133 of 136 (97.8 %) in the treatment group. On the morning of the third day a significantly lower Jackson score was seen in the treatment group (8.29 ± 5.07) compared to the control group (9.59 ± 5.31; p=0.035). This significant difference was maintained during day 4 and 5 becoming less evident on the morning of day 6 (p=0.056). The total Jackson score diminished below 1.0 on the morning of day 10 in the treatment group compared to the morning of day 12 in the control group.

The total Jackson score intensity over 12 days showed an AUC_1-12_ of 45.1 ± 32.5 compared to 53.8 ± 35.7 in the control group (p=0.023). The between group AUC_1-12_ differences for individual Jackson symptoms were consistently lower in the treatment group compared to the control group. The most prominent differences were seen in sore throat and blocked nose as shown in Figure 2 expressed as differences in AUC_1-12_ (95% CI). The mean daily sum score of morning and evening values are shown in Figure 3a, and scores of individual Jackson items over 12 days are shown in Appendix A.

**Figure 2:**
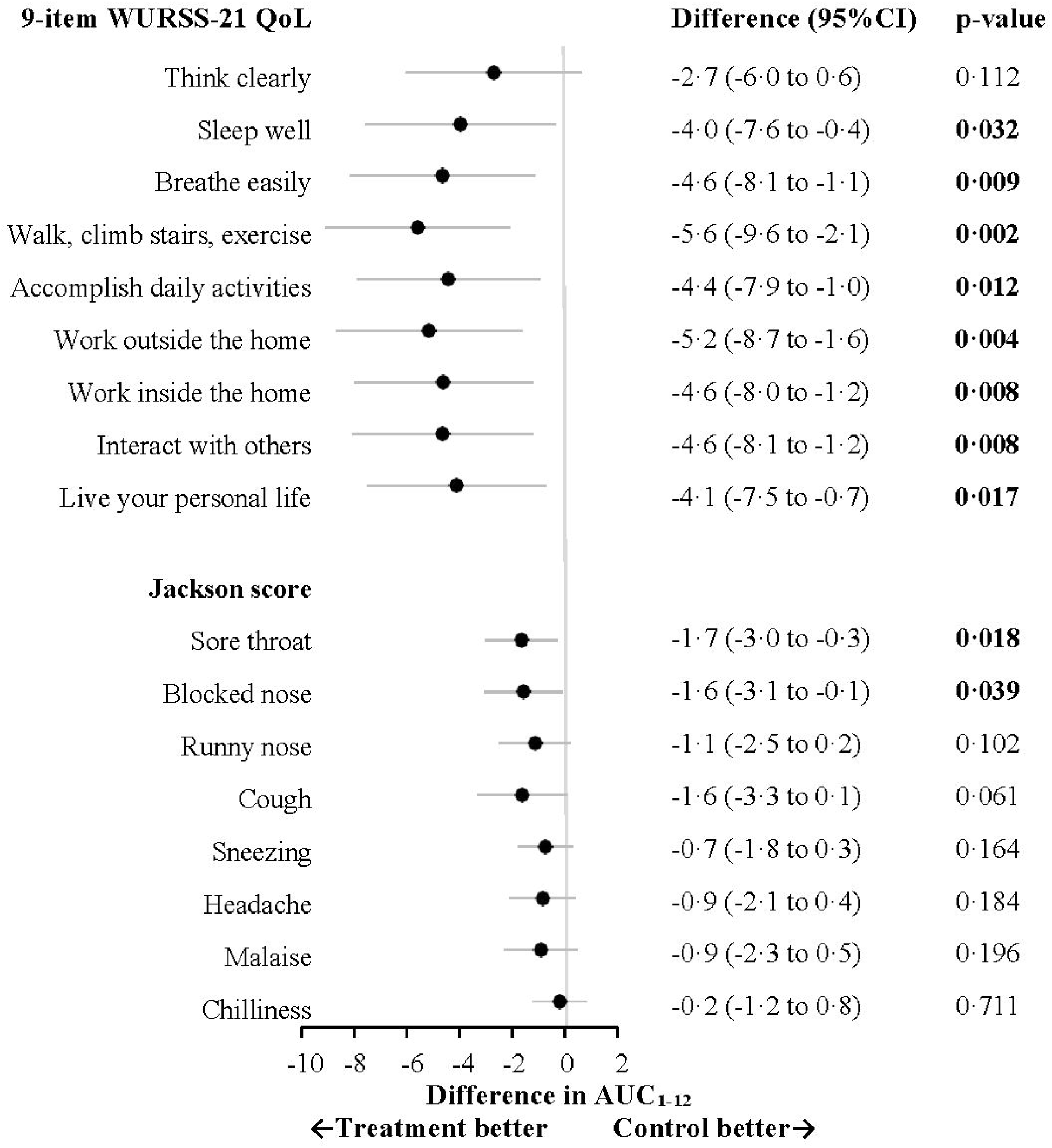
Forest plot analyses of individual items of 9-item WURSS-21 QoL domain and 8-item Jackson symptom ratings; significant differences shown in bold (p<0.05) WURSS=Wisconsin Upper Respiratory Symptom Survey, QoL=quality of life

**Figure 3:**
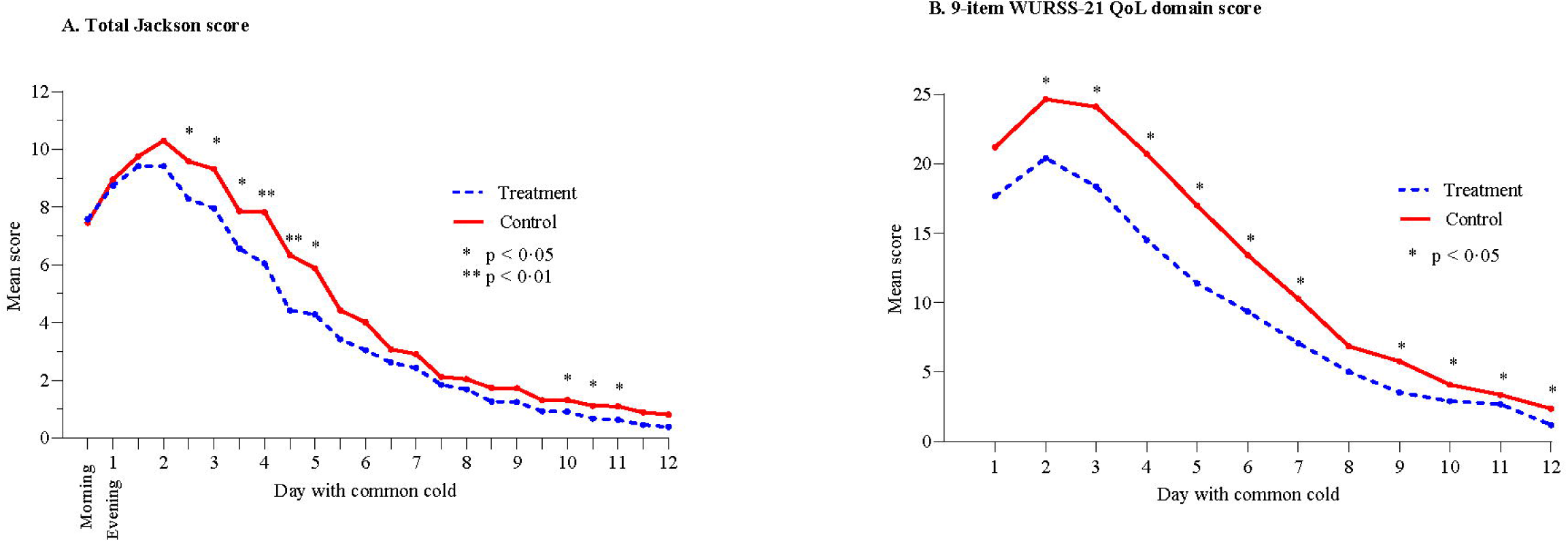
Mean daily sum score over 12 days based on morning and evening values for Jackson score and evening values for the 9-item WURSS-21 QoL domain. WURSS=Wisconsin Upper Respiratory Symptom Survey, QoL=quality of life.

One subject in the treatment group did not answer the 9-item WURSS-21 QoL domain questionnaire. Another 14 subjects (eight in the treatment group) misinterpreted the questionnaire by reporting a severe impact on quality of life at the end of the cold in spite of being fully recovered. Data from these subjects were considered invalid resulting in a FAS analysis of the 9-item WURSS-21 QoL domain of 127 and 125 subjects in the treatment and control group, respectively. The impact on quality of life integrated over 12 days was significantly lower in the treatment group with an AUC_1-12_ of 113.6 ± 107.7 compared to 152.7 ± 126.3 in the control group (p=0.006). The between group AUC_1-12_ differences were significantly lower for all individual items of the WURSS-21 QoL domain (Figure 2) in favour of treatment. The mean daily sum score of 9-item WURSS-21 QoL domain also showed a significant difference in favour of treatment on day two and thereafter (Figure 3b).

All nine individual QoL domain items were consistently lower with treatment compared to without treatment during the first week, with a significant difference seen within the first 12 hours for item 15 (walk, climb stairs, exercise) and item 19 (interact with others), as shown in Appendix B.

The duration of the common cold defined as the number of days from symptom start until the last day before answering “No” to the question “Do you think that you still have a common cold” for two days in a row was 6.26 ± 3.27 in the treatment group and 7.10 ± 4.06 in the control group (p=0.125). When defined as number of days with a total Jackson score > zero, colds duration in the treatment group was 7.11 ± 3.37 and in the control group 8.08 ± 4.13 (p=0.071). Corresponding data for the 9-item WURSS-21 QoL domain was 6.46 ± 3.48 and 7.62 ± 4.18 days respectively (p=0.030). The number of subjects scoring zero for individual Jackson symptoms day by day during the course of the common cold is shown in Table 2 and for the respective 9 items of WURSS-21 QoL domain in Table 3.

**Table 2:**
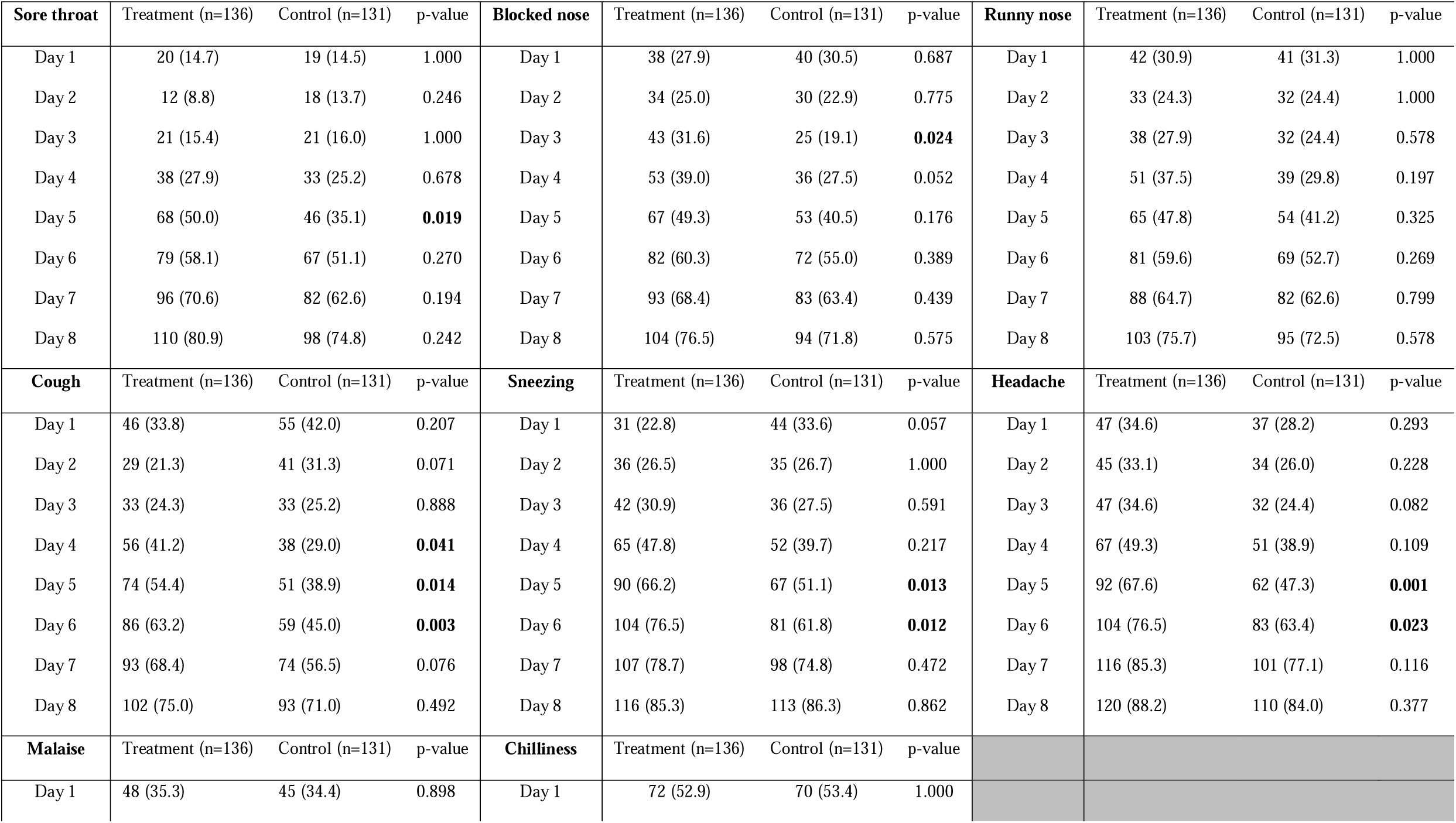
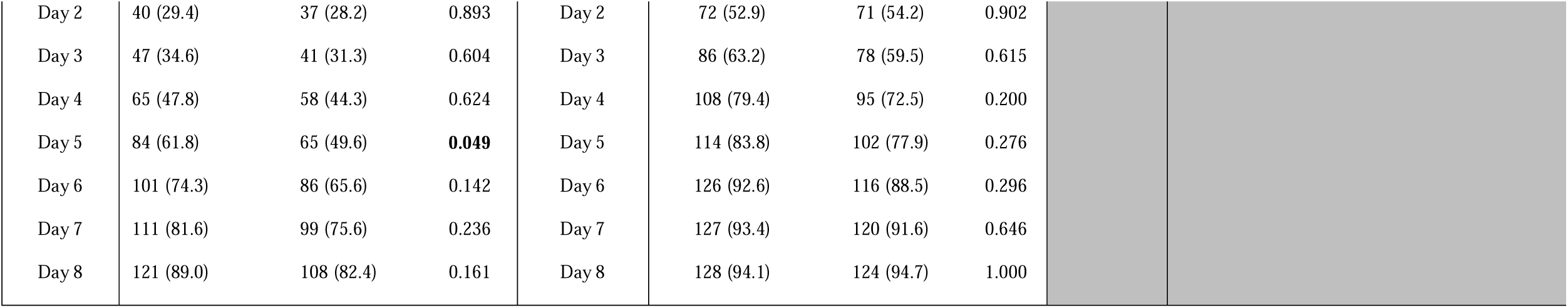
Number of subjects (% of subjects in respective group) without symptoms for each individual item of the Jackson score during the first 8 days of the common cold (mean of morning and evening ratings); bold values indicate significance below p=0.05.

| **Sore throat** | Treatment (n=136) | Control (n=131) | p-value | **Blocked nose** | Treatment (n=136) | Control (n=131) | p-value | **Runny nose** | Treatment (n=136) | Control (n=131) | p-value |
| --- | --- | --- | --- | --- | --- | --- | --- | --- | --- | --- | --- |
| Day 1 | 20 (14.7) | 19 (14.5) | 1.000 | Day 1 | 38 (27.9) | 40 (30.5) | 0.687 | Day 1 | 42 (30.9) | 41 (31.3) | 1.000 |
| Day 2 | 12 (8.8) | 18 (13.7) | 0.246 | Day 2 | 34 (25.0) | 30 (22.9) | 0.775 | Day 2 | 33 (24.3) | 32 (24.4) | 1.000 |
| Day 3 | 21 (15.4) | 21 (16.0) | 1.000 | Day 3 | 43 (31.6) | 25 (19.1) | **0.024** | Day 3 | 38 (27.9) | 32 (24.4) | 0.578 |
| Day 4 | 38 (27.9) | 33 (25.2) | 0.678 | Day 4 | 53 (39.0) | 36 (27.5) | 0.052 | Day 4 | 51 (37.5) | 39 (29.8) | 0.197 |
| Day 5 | 68 (50.0) | 46 (35.1) | **0.019** | Day 5 | 67 (49.3) | 53 (40.5) | 0.176 | Day 5 | 65 (47.8) | 54 (41.2) | 0.325 |
| Day 6 | 79 (58.1) | 67 (51.1) | 0.270 | Day 6 | 82 (60.3) | 72 (55.0) | 0.389 | Day 6 | 81 (59.6) | 69 (52.7) | 0.269 |
| Day 7 | 96 (70.6) | 82 (62.6) | 0.194 | Day 7 | 93 (68.4) | 83 (63.4) | 0.439 | Day 7 | 88 (64.7) | 82 (62.6) | 0.799 |
| Day 8 | 110 (80.9) | 98 (74.8) | 0.242 | Day 8 | 104 (76.5) | 94 (71.8) | 0.575 | Day 8 | 103 (75.7) | 95 (72.5) | 0.578 |
| **Cough** | Treatment (n=136) | Control (n=131) | p-value | **Sneezing** | Treatment (n=136) | Control (n=131) | p-value | **Headache** | Treatment (n=136) | Control (n=131) | p-value |
| Day 1 | 46 (33.8) | 55 (42.0) | 0.207 | Day 1 | 31 (22.8) | 44 (33.6) | 0.057 | Day 1 | 47 (34.6) | 37 (28.2) | 0.293 |
| Day 2 | 29 (21.3) | 41 (31.3) | 0.071 | Day 2 | 36 (26.5) | 35 (26.7) | 1.000 | Day 2 | 45 (33.1) | 34 (26.0) | 0.228 |
| Day 3 | 33 (24.3) | 33 (25.2) | 0.888 | Day 3 | 42 (30.9) | 36 (27.5) | 0.591 | Day 3 | 47 (34.6) | 32 (24.4) | 0.082 |
| Day 4 | 56 (41.2) | 38 (29.0) | **0.041** | Day 4 | 65 (47.8) | 52 (39.7) | 0.217 | Day 4 | 67 (49.3) | 51 (38.9) | 0.109 |
| Day 5 | 74 (54.4) | 51 (38.9) | **0.014** | Day 5 | 90 (66.2) | 67 (51.1) | **0.013** | Day 5 | 92 (67.6) | 62 (47.3) | **0.001** |
| Day 6 | 86 (63.2) | 59 (45.0) | **0.003** | Day 6 | 104 (76.5) | 81 (61.8) | **0.012** | Day 6 | 104 (76.5) | 83 (63.4) | **0.023** |
| Day 7 | 93 (68.4) | 74 (56.5) | 0.076 | Day 7 | 107 (78.7) | 98 (74.8) | 0.472 | Day 7 | 116 (85.3) | 101 (77.1) | 0.116 |
| Day 8 | 102 (75.0) | 93 (71.0) | 0.492 | Day 8 | 116 (85.3) | 113 (86.3) | 0.862 | Day 8 | 120 (88.2) | 110 (84.0) | 0.377 |
| **Malaise** | Treatment (n=136) | Control (n=131) | p-value | **Chilliness** | Treatment (n=136) | Control (n=131) | p-value |  |  |  |  |
| Day 1 | 48 (35.3) | 45 (34.4) | 0.898 | Day 1 | 72 (52.9) | 70 (53.4) | 1.000 |  |  |  |  |

|  |  |  |  |  |  |  |  |
| --- | --- | --- | --- | --- | --- | --- | --- |
| Day 2 | 40 (29.4) | 37 (28.2) | 0.893 | Day 2 | 72 (52.9) | 71 (54.2) | 0.902 |
| Day 3 | 47 (34.6) | 41 (31.3) | 0.604 | Day 3 | 86 (63.2) | 78 (59.5) | 0.615 |
| Day 4 | 65 (47.8) | 58 (44.3) | 0.624 | Day 4 | 108 (79.4) | 95 (72.5) | 0.200 |
| Day 5 | 84 (61.8) | 65 (49.6) | **0.049** | Day 5 | 114 (83.8) | 102 (77.9) | 0.276 |
| Day 6 | 101 (74.3) | 86 (65.6) | 0.142 | Day 6 | 126 (92.6) | 116 (88.5) | 0.296 |
| Day 7 | 111 (81.6) | 99 (75.6) | 0.236 | Day 7 | 127 (93.4) | 120 (91.6) | 0.646 |
| Day 8 | 121 (89.0) | 108 (82.4) | 0.161 | Day 8 | 128 (94.1) | 124 (94.7) | 1.000 |

**Table 3:** Number of subjects (% of subjects in respective group) without symptoms for each individual item of the 9-item WURSS-21 QoL domain during the first 8 days of the common cold (evening ratings); bold values indicate significance below p=0.05.

| Item 12 Think clearly |  |  |  | Item 13 Sleep well |  |  |  | Item 14 Breathe easily |  |  |  |
| --- | --- | --- | --- | --- | --- | --- | --- | --- | --- | --- | --- |
| Item 12 | Treatment (n=127) | Control (n=125) | p-value | Item 13 | Treatment (n=127) | Control (n=125) | p-value | Item 14 | Treatment (n=127) | Control (n=125) | p-value |
| Day 1 | 63 (49.6) | 54 (43.2) | 0.316 | Day 1 | 29 (22.8) | 24 (19.2) | 0.538 | Day 1 | 20 (15.7) | 22 (17.6) | 0.737 |
| Day 2 | 51 (40.2) | 34 (27.2) | **0.033** | Day 2 | 21 (16.5) | 12 (9.6) | 0.135 | Day 2 | 18 (14.2) | 15 (12.0) | 0.710 |
| Day 3 | 58 (45.7) | 37 (29.6) | **0.009** | Day 3 | 28 (22.0) | 21 (16.8) | 0.341 | Day 3 | 26 (20.5) | 18 (14.4) | 0.246 |
| Day 4 | 68 (53.5) | 44 (35.2) | **0.002** | Day 4 | 40 (31.5) | 27 (21.6) | 0.088 | Day 4 | 42 (33.1) | 28 (22.4) | 0.068 |
| Day 5 | 83 (65.4) | 52 (41.6) | **<0.001** | Day 5 | 57 (44.9) | 38 (30.4) | **0.020** | Day 5 | 55 (43.3) | 39 (31.2) | 0.052 |
| Day 6 | 90 (70.9) | 65 (52.0) | **0.003** | Day 6 | 69 (54.3) | 47 (37.6) | **0.008** | Day 6 | 61 (48.0) | 47 (37.6) | 0.100 |
| Day 7 | 98 (77.2) | 84 (67.2) | 0.092 | Day 7 | 83 (65.4) | 61 (48.8) | **0.011** | Day 7 | 80 (63.0) | 59 (47.2) | **0.016** |
| Day 8 | 106 (83.5) | 92 (73.6) | 0.066 | Day 8 | 95 (74.8) | 78 (62.4) | **0.042** | Day 8 | 89 (70.1) | 78 (62.4) | 0.231 |
| Item 15 Walk, climb stairs, exercise |  |  |  | Item 16 Accomplish daily activities |  |  |  | Item 17 Work outside the home |  |  |  |
| Item 15 | Treatment (n=127) | Control (n=125) | p-value | Item 16 | Treatment (n=127) | Control (n=125) | p-value | Item 17 | Treatment (n=127) | Control (n=125) | p-value |
| Day 1 | 48 (37.8) | 36 (28.8) | 0.143 | Day 1 | 46 (36.2) | 36 (28.8) | 0.228 | Day 1 | 50 (39.4) | 39 (31.2) | 0.189 |
| Day 2 | 37 (29.1) | 26 (20.8) | 0.146 | Day 2 | 33 (26.0) | 26 (20.8) | 0.373 | Day 2 | 41 (32.3) | 24 (19.2) | 0.021 |
| Day 3 | 44 (34.6) | 29 (23.2) | 0.052 | Day 3 | 43 (33.9) | 29 (23.2) | 0.070 | Day 3 | 47 (37.0) | 34 (27.2) | 0.107 |
| Day 4 | 59 (46.5) | 31 (24.8) | **<0.001** | Day 4 | 63 (49.6) | 36 (28.8) | **0.001** | Day 4 | 61 (48.0) | 38 (30.4) | **0.005** |
| Day 5 | 70 (55.1) | 43 (34.4) | **0.001** | Day 5 | 77 (60.6) | 50 (40.0) | **0.002** | Day 5 | 77 (60.6) | 49 (39.2) | **0.001** |
| Day 6 | 84 (66.1) | 55 (44.4) | **0.001** | Day 6 | 89 (70.1) | 58 (46.4) | **<0.001** | Day 6 | 91 (71.7) | 60 (48.0) | **<0.001** |
| Day 7 | 95 (74.8) | 73 (58.4) | **0.007** | Day 7 | 97 (76.4) | 78 (62.4) | **0.020** | Day 7 | 97 (76.4) | 76 (60.8) | **0.010** |
| Day 8 | 104 (81.9) | 89 (71.2) | 0.053 | Day 8 | 106 (83.5) | 93 (74.4) | 0.090 | Day 8 | 107 (84.3) | 91 (72.8) | **0.032** |
| Item 18 Work inside the home |  |  |  | Item 19 Interact with others |  |  |  | Item 20 Live your personal life |  |  |  |
| Item 18 | Treatment (n=127) | Control (n=125) | p-value | Item 19 | Treatment (n=127) | Control (n=125) | p-value | Item 20 | Treatment (n=127) | Control (n=125) | p-value |
| Day 1 | 51 (40.2) | 40 (32.0) | 0.192 | Day 1 | 49 (38.6) | 42 (33.6) | 0.433 | Day 1 | 48 (37.8) | 43 (34.4) | 0.602 |
| Day 2 | 40 (31.5) | 28 (22.4) | 0.119 | Day 2 | 51 (40.2) | 34 (27.2) | 0.033 | Day 2 | 43 (33.9) | 33 (26.4) | 0.218 |
| Day 3 | 46 (36.2) | 34 (27.2) | 0.138 | Day 3 | 51 (40.2) | 35 (28.0) | 0.047 | Day 3 | 53 (41.7) | 35 (28.0) | **0.025** |
| Day 4 | 66 (52.0) | 41 (32.8) | **0.002** | Day 4 | 67 (52.8) | 41 (32.8) | **0.002** | Day 4 | 68 (53.6) | 42 (33.6) | **0.002** |
| Day 5 | 82 (64.6) | 55 (44.0) | **0.002** | Day 5 | 77 (60.6) | 55 (44.0) | **0.011** | Day 5 | 83 (65.4) | 56 (44.8) | **0.001** |
| Day 6 | 91 (71.7) | 62 (49.6) | **<0.001** | Day 6 | 90 (70.9) | 65 (52.0) | **0.003** | Day 6 | 92 (72.4) | 71 (56.8) | **0.012** |
| Day 7 | 99 (78.0) | 80 (64.0) | **0.018** | Day 7 | 95 (74.8) | 83 (66.4) | 0.167 | Day 7 | 98 (77.2) | 88 (70.4) | 0.253 |
| Day 8 | 109 (85.8) | 95 (76.0) | 0.055 | Day 8 | 106 (83.5) | 95 (76.0) | 0.160 | Day 8 | 108 (85.0) | 98 (78.4) | 0.194 |

WURSS=Wisconsin Upper Respiratory Symptom Survey, QoL= Quality of Life

Concomitant medication was used less often (2.3 ± 3.0 days) by the treatment group than the control group (3.2 ± 3.2 days, p=0.032) over the first 7 days of a cold and over 14 days (5.0 ± 6.4 versus 6.7 ± 6.7, p=0.032). During the first week a significant difference in the number of subjects using symptom relieving medications was seen on day one, three and four, as shown in Figure 4.

**Figure 4:**
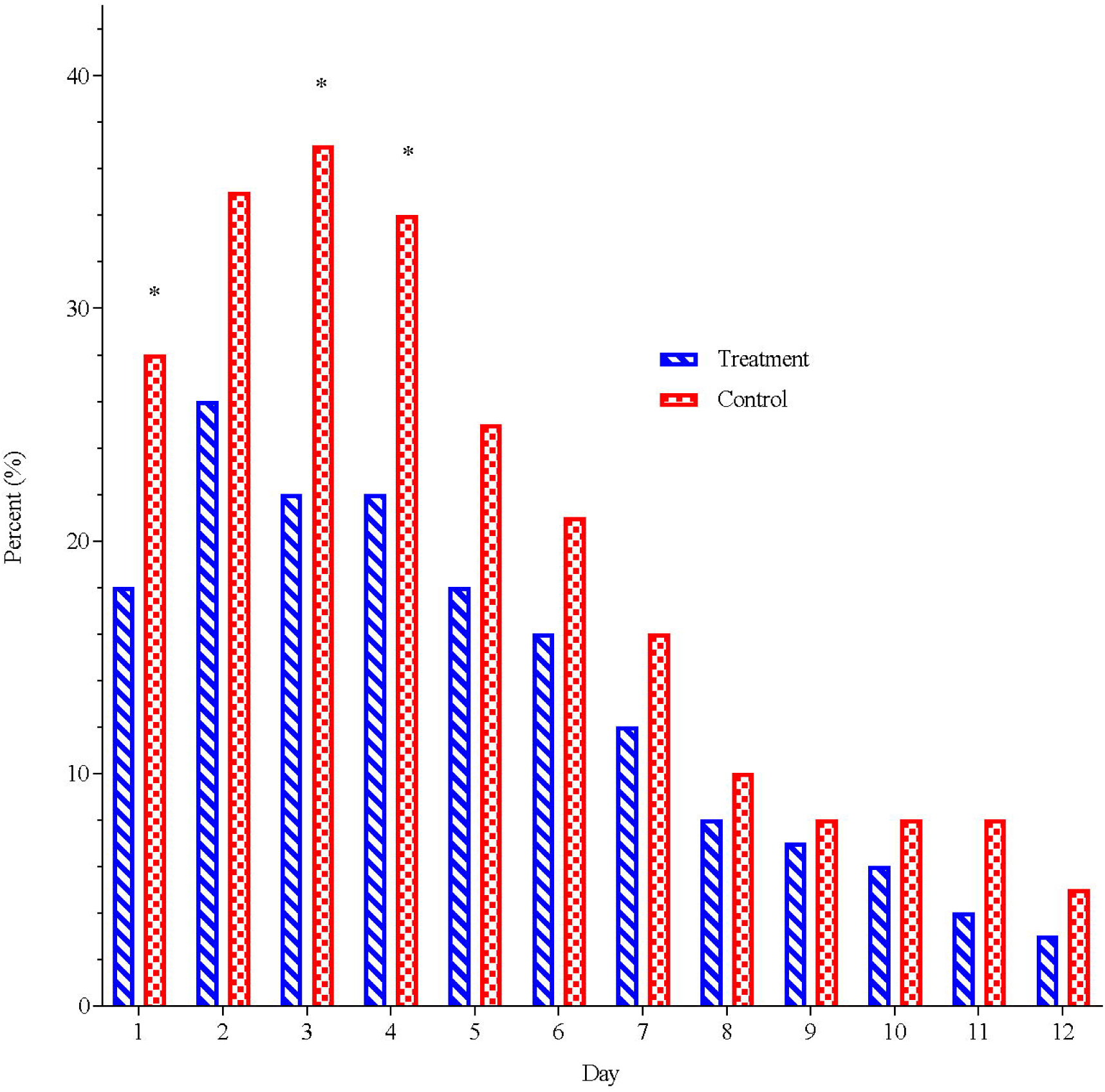
Daily frequency of subjects using rescue medication. * p<0.05

Of the 136 participants in the treatment group 115 evaluated the efficacy at the end of the study. The assessment “very good” or “good” was admitted by 95 subjects (82.6%). Product tolerability and safety was evaluated by 116 subjects and was considered to be “very good” or “good” by 115 (99.1%) and 114 (98.3%), respectively. A total of 14 subjects (14 of 400; 3.5%) suffered from adverse events, four in the treatment group compared to ten in the control group. No adverse event was considered by the investigators to be related to treatment. One serious adverse event, appendicitis, occurred in the treatment group.

## CONCLUSION

This single-blinded, randomised controlled trial was conducted to assess whether this study design can adequately evaluate the common cold using symptom scoring with the Jackson scale as well as by quality of life scoring with the 9-item WURSS-21 QoL domain. A secondary objective was to investigate whether treatment with a glycerol throat spray containing cold-adapted cod trypsin (GCTS) when initiated at the first self-perceived prodromal symptoms can alleviate and shorten a common cold. The 9-item WURSS-21 QoL domain composite score, as well as eight out of nine individual QoL items, seem to be a most sensitive instrument for demonstrating that GCTS treatment compared to no treatment significantly improves the colds sufferers’ quality of life over 12 days (AUC_1-12_). The reduction in Jackson AUC_1-12_ was likewise significantly reduced with active treatment although less discriminating than the 9-item WURSS-21 QoL domain. These results, together with significantly less use of rescue medication, and an approximately one day quicker recovery are all indicative of a positive effect after treatment with a glycerol-cod trypsin throat spray (GCTS).

The outcome of this study supports previous findings of a randomised placebo-controlled pilot study on healthy volunteers treated with the same glycerol-cod trypsin throat spray in which a lower oropharyngeal virus load and a reduced duration of common cold symptoms was shown following experimental intranasal inoculation of rhinovirus-16 [12]. Further, two open-label studies using participants as their own historical controls showed reduced sick leave time in kindergarten employees [17] and elderly care personnel [18] when treated with the glycerol-cod trypsin spray during the common cold season.

Interestingly, the 9 item WURSS-21 QoL domain was more informative than the total Jackson symptom severity scores in discerning the effect of treatment particularly on days 1 and 2. The 9 item WURSS-21 QoL domain found significant differences benefiting the treated group beginning one day earlier (day 2 vs. day 3) compared to the Jackson scale. It might be argued that this early difference in quality of life is the result of inclusion skewness nonetheless randomisation and an almost identical Jackson score from the start favour an early alleviating effect of treatment best detected by the WURSS-21 QoL domain.

Many, if not most, prior experimental protocol designs have relied on the Jackson scale [13] for self-diagnosis at entry and to describe the evolution of symptoms despite the fact that it is not validated and often modified. Nonetheless, most studies evaluating treatments for the common cold have employed Jackson criteria as a primary outcome measure [4, 19] or for validating new questionnaires [14, 20], or assessing the relationship between common cold symptoms and biological parameters [21]. On the contrary, the WURSS-21 questionnaire [14] is an illness-specific health-related quality of life instrument that has been validated against the Jackson scale and the SF-8 Health Survey, demonstrating a high correlation. The 9-item quality of life domain of WURSS-21 has not been validated separately but does show a correlation with Jackson score in the present study (see Appendix C). Although the correlation is not perfect, a Bland-Altman analysis shows that the limits of agreement, a measure of both bias and random variation, correspond to two to three times what is generally considered a minimal clinically important difference of WURSS-21, i.e. 1 unit of 7.

Furthermore, the differences between the two scores seem to be mainly random, reflecting the scores reliability rather than bias.

Investigations into the sources of bias in the design of randomised clinical trials have found that binary variables are generally less subject to bias than continuous variables [22]. In this design all subjects made a binary decision as to when they became ill with a common cold (the start) and made subsequent daily binary decisions supporting how long they were ill.

Thus, while these decisions must integrate internal signals of the state of their illness, they are nevertheless binary determinants of illness presence and duration. Using these measures, the treatment group demonstrated a significant difference from the no treatment group in illness duration using the QoL instrument (p=0.03) and near significance using the Jackson scale to determine number of days greater than zero (p=0.07). These results accord with overall efficacy outcomes derived from subjective symptom and QoL scales including overall symptom burdens (AUCs).

According to Krogsbøll et al [23], in designs with a treatment, placebo and no treatment arms there are three sources of positive effect: placebo, treatment and spontaneous improvement. The latter describes all acute self-limiting conditions including the common cold. In this design the latter would be accelerated by palliative treatment. The no treatment group acts as a control for spontaneous improvement. The placebo effect is generally driven by both expectation and conditioning [24]. In these two respects consumers would not expect oral sprays to be effective as treatments for a nasal disorder, but rather nasal saline, nasal decongestants, or colds tablets. Lastly, in normal subjects without preconditioning, no placebo effect (lactose tablet) was found over no treatment for subjective expectancy, or their outcome measures, using a battery of 5 psychological tests sensitive to biases in emotional processing that occur prior to subjective change [25].

There are possible limitations to this study. First, participants were asked to self-diagnose the onset of an emerging common cold while simultaneously experiencing a total Jackson score of at least one for any symptom (except headache). Higher Jackson scores are normally required to accept inclusion in symptomatic relief trials [19, 26]. But to reduce early virus propagation and replication it was judged essential that the tested throat spray is applied as early as possible. Since no objective signs of a common cold are present during prodrome, we instead hypothesized that years of personal experience catching colds makes each subject the best predictor of their imminent illness. Such methods are similar to those of other natural colds studies [8, 14 and 26].

Second, while the primary objective of this methods trial was to assess the suitability of different rating instruments to detect illness burden and treatment effects this study was secondarily designed to provide a proof of concept of the tested product. Despite the fact that multiple endpoints were studied it is unlikely that these positive results were caused by chance due to multiplicity since pre-defined hypotheses were tested and the multiple endpoints were not independent but related. The consistent results in favour of active treatment point towards a true effect of the tested product on colds illness.

Third, since this trial was designed to compare active treatment with a medical device to no treatment it is possible that the observed outcomes were due to a placebo effect. The no treatment group should adequately control for a self-limiting condition or spontaneous improvement [27] in a randomised clinical trial with patient-reported outcomes [28]. More recently a study looking at the placebo effect of oral *Echinacea* for treatment of the common cold, using *a priori* analyses of cold duration and the AUC for the WURSS-21, determined that the placebo effect was limited [29].

The results presented here indicate that a quality of life instrument (9-item WURSS-21 QoL domain) can detect colds illness and the effect of treatment with a throat spray designed as an active physical barrier to viral entry into mucosal cells in the oro-pharynx [7, 9–11].

In summary, the results of this methods trial remind us that consumers integrate the effects of emerging symptom complexes and quickly recognize the signals of a common cold illness and their emerging reduced functionality. Given heightened global awareness of other circulating respiratory viruses of greater morbidity, such as SARS-CoV-2 and influenza, quality of life domains should be considered in determining wellness and recovery from these infections.

## Data Availability

Individual participant data, after having been anonymized will be made available, along with the study protocol, statistical analysis plan and consent form. Data will be made available beginning 3 months and ending 5 years after publication of the article to researchers whose proposed use of the data is approved by the original study investigators. Proposals should be directed to the corresponding author and requesters will need to sign a data access agreement.

## Appendices

Appendix A: Jackson score individual items morning and evening day 1-12

Appendix B: 9-item WURSS-21 QoL domain individual items: Daily mean score day 1-12

Appendix C: Comparison between Jackson score and 9-item WURSS-21 QoL domain

