## Supplementary figures and images for "Early intervention with a glycerol throat spray containing cold-adapted cod trypsin after self-diagnosis of the common cold: a randomised prospective, parallel group and single-blind methods trial (Glycerol-cod trypsin spray in common cold)"

### Appendix A

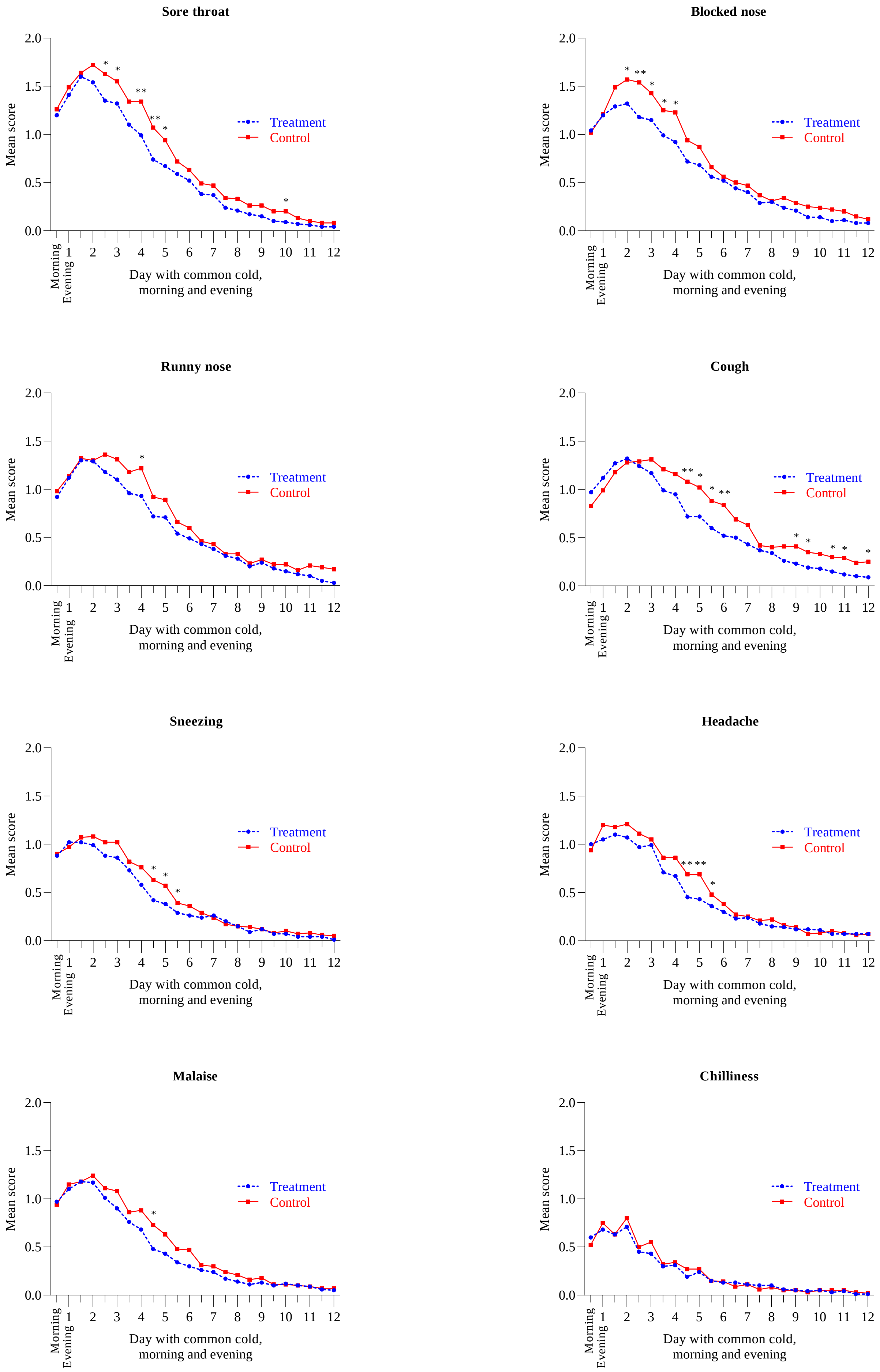


***Figure A:*** **Jackson score individual items morning and evening day 1-12**

### Appendix B

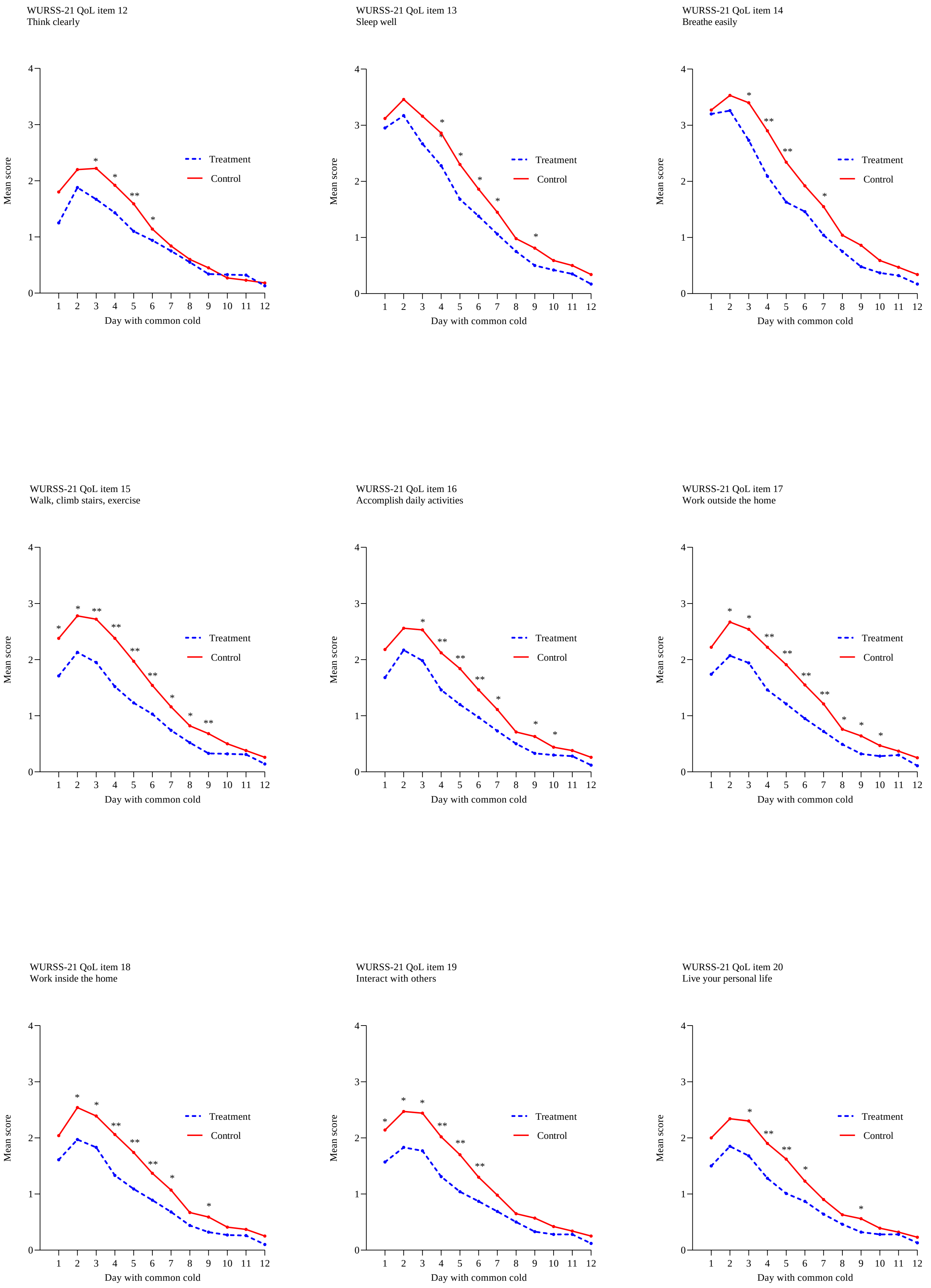


***Figure B:*** **9-item WURSS-21 QoL domain individual items: Daily mean score day 1-12**

### Appendix C

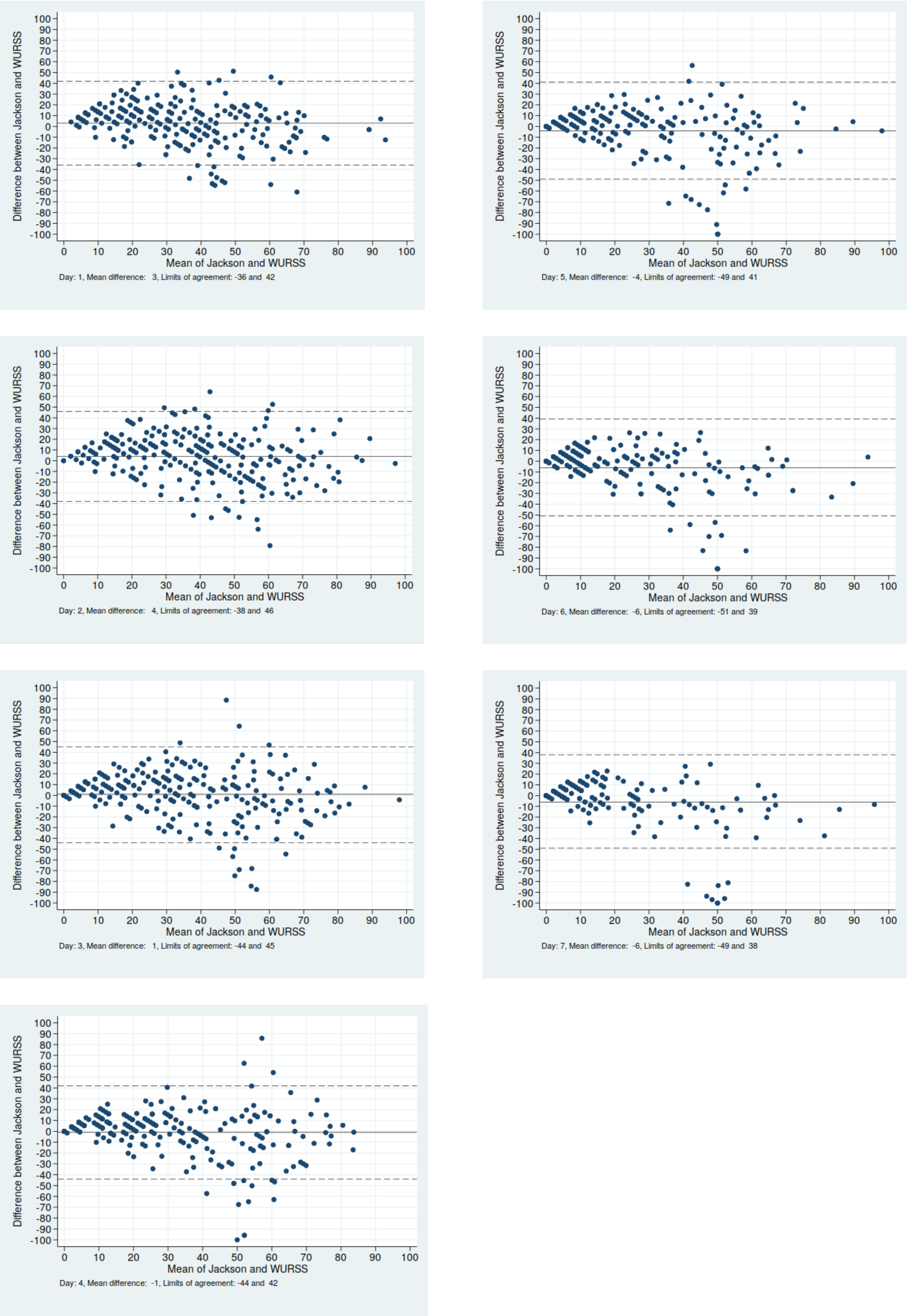


***Figure C:* Comparison between Jackson score (evening) and 9-item WURSS-21 QoL domain day 1-7**
